# Predicting outcome after newborn stroke: A lesion network mapping study leveraging large-scale data

**DOI:** 10.1101/2025.08.26.25334501

**Authors:** Claire E. Kelly, Jian Chen ME, Richard Beare, Belinda Stojanovskix, Jesse S. Shapiro, Sebastian Grunt, Nedelina Slavova, Manuela Pastore-Wapp, Maja I. Steinlin, Mark T. Mackay, Joseph Y.M. Yang

## Abstract

**Background:** Predicting development of cerebral palsy following neonatal stroke remains challenging. This study aimed to identify novel acute brain functional connectome-based correlates of cerebral palsy following neonatal stroke.

**Methods:** Stroke lesions were segmented from routine clinical diffusion images of a cohort of term-born neonates with symptomatic arterial ischemic stroke, recruited to Swiss and Australian pediatric stroke registries. Lesions, and 3T resting state functional MRIs of term-born newborns from the developing Human Connectome Project, were co-registered to a template. A neonatal stroke functional connectome was created by computing voxel-wise correlations between lesions and gray matter regions. Linear regressions compared functional connections to lesions between participants who did and did not develop cerebral palsy.

**Results:** Eighty-five newborns with stroke were included (64% male; median age at MRI of 4 days), of which 33% developed cerebral palsy at a median age of 2.1 years. Multiple gray matter regions were more highly functionally correlated to lesions in participants who developed cerebral palsy (1721 voxels; *t*: 5.4-7.4; all *p*<0.05, family-wise error rate corrected). These regions included the basal ganglia, thalamus, cerebellum, frontal regions (inferior and orbital frontal and superior frontal), temporal regions (pole, superior, and mesial temporal including hippocampus and amygdala) and the insula.

**Conclusions:** This study identified functional networks related to the development of cerebral palsy following neonatal stroke. Building on prior individual lesion-based studies, this work suggests that development of cerebral palsy after neonatal stroke is related to disruptions of broader functional networks involving motor and extramotor regions, as opposed to only lesions in motor regions.

## Introduction

The perinatal period involves both dynamic brain growth and vulnerability to brain injury, including neonatal arterial ischemic stroke (NAIS).^1–4^ NAIS is associated with lifelong disabilities, such as cerebral palsy (over one third of cases).^4^ Prior work often predicted development of cerebral palsy by analyzing characteristics of individual brain lesions, using qualitative or quantitative region-based or voxel-based approaches, such as voxel-based lesion-symptom mapping (VLSM).^5–12^ Disruptions to primary motor regions and tracts were related to the development of cerebral palsy.^5–12^ However, lesion-based analyses have not fully explained motor outcome following NAIS.^5^

Development of cerebral palsy may be more fully explained by network dysfunction beyond lesions. Network dysfunction could occur through functional disconnection or diaschisis (physiological changes in areas distant from, but functionally connected to, stroke lesions).^13,14^ These phenomena are known to occur in adults.^13^ In NAIS, lesion-based studies have shown that lesions affect primary motor regions, but imaging changes can also be seen outside primary motor regions (e.g., the thalamus, basal ganglia, and corpus callosum).^5, 14^ This provides initial support for the idea that widespread network disruptions occur in the newborn period after stroke.^5, 14^ Evidence for network dysfunction after NAIS, however, remains limited by a lack of systematic investigations using specific network-based newborn neuroimaging techniques in representative cohorts.

One approach for studying brain connectivity is to use resting-state functional MRI (fMRI) scans, which measure spontaneous fluctuations in blood oxygenation level, an indirect marker of neuronal activity.^13, 15^ When activity of different regions is correlated, the regions are considered functionally connected as part of networks.^13^ A body of work has documented multiple newborn resting-state networks.^16–19^ Acquisition of resting-state scans for neonates with stroke is time consuming, expensive, and difficult, if not infeasible, to implement clinically.^17^

Lesion network mapping was developed with the goal of measuring brain connectivity following injuries in ways previously considered infeasible.^13^ Lesion network mapping requires only the collection of routine, clinical MRIs from patients, while specialized connectome information is derived from more readily available datasets.^13^ One such dataset is the Human Connectome Project, where resting-state fMRI scans were acquired from thousands of adults, mapping the normal functional connectome (the collective ensemble of functional connections in the brain).^20^ More recently, the developing Human Connectome Project collected resting-state scans for hundreds of newborns.^19^ Through combining lesion location from clinical images with connectome information from large-scale datasets, lesion network mapping derives functional connections to stroke lesions.^13^ This output of lesion network mapping can be considered the stroke functional connectome (or disconnectome, i.e., gray matter regions forming disrupted functional networks following stroke).

Lesion network mapping has been extensively applied and compared against direct connectivity methods in adults.^21–23^ Lesion network mapping may provide new neuroimaging predictors of cerebral palsy, but this technique has not yet been applied in NAIS. Our aim was to identify novel neuroimaging correlates of cerebral palsy, through leveraging clinically indicated acute diagnostic MRIs of newborns with symptomatic NAIS, and normative resting-state fMRI data from the developing Human Connectome Project. We hypothesized that the neonatal stroke functional connectome is related to development of cerebral palsy.

## Methods

### Reporting guideline

The STROBE guidelines for observational studies were followed.

### Data availability statement

Data for the developing Human Connectome Project are available on the NIMH Data Archive. Instructions on how to access these data are available here: https://biomedia.github.io/dHCP-release-notes/. Anonymized data for the newborn stroke cohorts can be shared upon appropriate request, subject to ethical approval and adequate data sharing agreements.

### Participants and procedures

#### Patients with NAIS

This research was based on two representative cohorts of newborns with NAIS recruited to pediatric stroke registries in Australia and Switzerland.^5, 9, 24^ All key information about these cohorts relevant to the current manuscript are reported in the methods and results. Participants were newborns who were born at term (>36 weeks of gestation) and had radiological evidence of acute infarction on neonatal MRI. Newborns with subacute infarction, other injuries or dual pathology were excluded, including hypoxic ischemic encephalopathy, external border-zone infarction, or hemorrhagic stroke.^5, 9, 24^ Neonatal MRI scans, including T2-weighted and diffusion imaging sequences, were acquired during routine clinical care from a range of clinical sites, where uniform scanner type, field strength and acquisition parameters were not possible. Clinical outcomes were determined at a minimum of 18 months after neonatal stroke. Cerebral palsy was diagnosed based on clinical examination performed by a neurologist or pediatrician, according to guidelines of the Surveillance Group of Cerebral Palsy in Europe,^25^ and severity was classified with the Gross Motor Functional Classification System.^26^ Data from a total of 85 newborns with NAIS were included in the current study (participant numbers are described in more detail in the Results). This study was approved by the Royal Children’s Hospital Melbourne Human Research Ethics Committee (HREC 37115). The Swiss Neuropediatric Stroke Registry was approved by the local ethics committee in Bern, and informed consent was obtained from the parents of Swiss children with NAIS.^5, 9, 24^

#### Large-scale, normative neuroimaging dataset

The current study also utilized data from the developing Human Connectome Project cohort.^19, 27, 28^ Data were downloaded from the developing Human Connectome Project’s third open access data release. This release consisted of multi-modal MRI scans and key demographic data (including age at birth, birth weight, and sex). Anatomical T2-weighted images and resting state fMRI images were required and downloaded for the current study. These MRI data that were downloaded were already minimally pre-processed MRI data, rather than raw MRI data. We selected MRI and demographic data for live newborn infants born at term and scanned at term age (37-44 weeks).^19, 27, 28^ This age at birth and scanning meant that the data were appropriate to utilize in the current study as a normative reference for the term-born NAIS cohort.

Regarding MRI acquisition, the MRI scanning was conducted on a 3-Tesla (3T) Philips Achieva scanner using a dedicated neonatal imaging system located at the Evelina Newborn Imaging Centre, St Thomas’ Hospital, London, UK. T2-weighted images were acquired in sagittal and axial slice stacks with in-plane resolution of 0.8 x 0.8 mm^2^ and 1.6 mm slices overlapped by 0.8 mm, 12000 ms repetition time (TR), 156 ms echo time (TE), and sensitivity encoding (SENSE) factor 2.11 (axial) and 2.60 (sagittal). Resting-state fMRI images were acquired using a high temporal resolution sequence developed for neonates, with multiband 9x accelerated echo-planar imaging, acquisition time 15 minutes, 38 ms TE, 392 ms TR, 2300 volumes, and an acquired resolution of 2.15 mm isotropic.^29^ Along with the functional images, additional spin-echo acquisitions with both anterior-posterior (AP) and posterior-anterior (PA) encoding directions were also acquired.

Regarding the MRI minimal pre-processing, a template image was created from the 40-week T2-weighted images and provided with the data release.^30^ The minimal pre-processing of the functional images included slice-to-volume motion correction, dynamic susceptibility distortion correction making use of the spin-echo images with opposing phase-encoding directions, and independent component analysis (ICA)-based denoising.^31^ Global (whole brain) signal regression was not performed prior to or after the data download.^31^ The functional images were registered to the developing Human Connectome Project 40-week T2-weighted template, and transformations between the pre-processed functional images and the template were provided in the data release.^31^

Stringent quality control of all raw and derived imaging data was performed both prior to the data release, and additionally after downloading the data during all reported image analysis steps. Any poor-quality imaging data were excluded, meaning data from a total of 518 normative newborns were included in the current study (participant numbers are described in more detail in the Results section).

The developing Human Connectome Project study was approved by the UK Health Research Authority (14/LO/1169). Written parental consent was obtained in every case for imaging and data release.

### Brain imaging analysis: Creating the neonatal stroke functional connectome

Making use of both the NAIS and developing Human Connectome Project data, we developed and applied a novel neonatal lesion network mapping approach (Fig 1). Our pipeline, performed by experienced image analysts, can be broken down into four major components: 1) Delineation of neonatal stroke lesions; 2) Establishment of the normative neonatal functional connectome; 3) Computing the functional connections to stroke lesions; 4) Statistical analysis. All key methodological details relevant to the current manuscript are described in the sections below.

**Fig 1.**
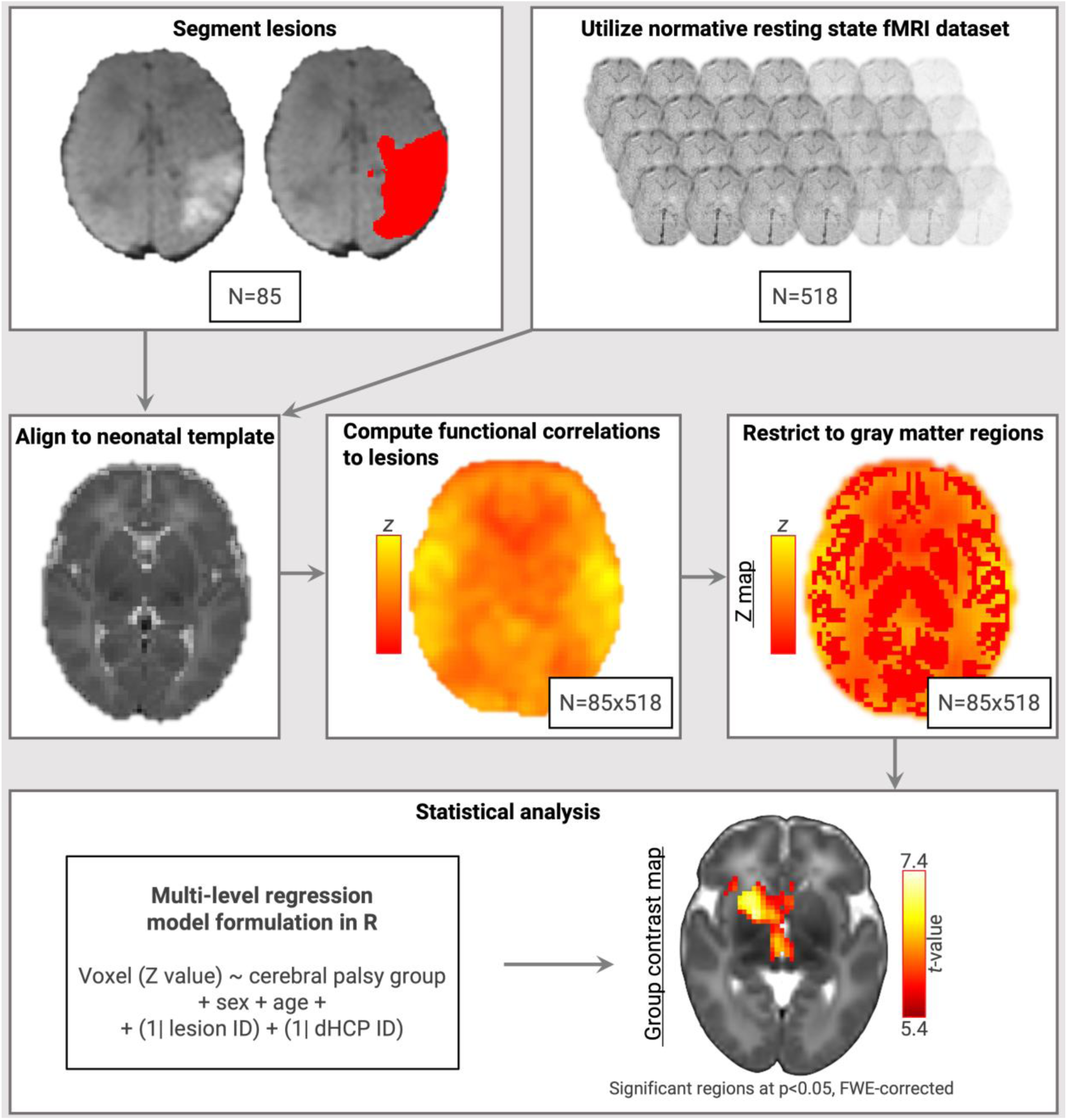
Overview of the novel neonatal lesion network mapping framework developed and applied in the current study. dHCP = developing Human Connectome Project; ID = identifier; fMRI = functional magnetic resonance imaging; FWE = family-wise error rate-corrected; R = the R software used for statistical analyses; Z = correlation coefficient (for the functional correlation between lesions and other brain regions) transformed to a z distribution. All steps shown are described in detail in the methods section of the current study. The model formulation shown (bottom left) is described in detail in the methods section of the current study (sub-section “Statistical analysis”).

#### Delineation of neonatal stroke lesions

Stroke lesions were manually segmented slice-by-slice on the diffusion MRI scans of the neonates with NAIS by a pediatric neurologist.^5^ As shown in our prior work based on the same cohort, the brain regions affected by stroke lesions were primarily located in unilateral middle cerebral artery (MCA) territory and encompassed gray matter and white matter but were primarily gray matter based, which is consistent with prior findings in other cohorts.^5, 12^ The resulting stroke lesion mask images in diffusion image space were transformed to their corresponding T2-weighted structural image space and then to the space of a common template image; the developing Human Connectome Project 40-week T2-weighted template. Transformations were performed using a combination of linear warping [with the Functional MRI of the Brain Software Library (FSL) Linear Image Registration Tool (FLIRT)^32–34^] and affine nonlinear warping (with the Advanced Normalization Tools (ANTs) software package^35^).

The stroke lesion masks were kept in their original brain hemisphere (left or right hemisphere). The lesions did not need to be projected to the one hemisphere for the current connectome analysis (i.e., this is different to lesion mask processing steps in past VLSM studies of NAIS populations, where lesions had to be projected to the same (left) hemisphere^5^).

#### The normative neonatal functional connectome

We aligned the minimally pre-processed resting-state fMRI images of each of the 518 neonates from the developing Human Connectome Project with the common template image (the developing Human Connectome Project 40-week T2-weighted template). This alignment made use of the transformation matrices already provided in the developing Human Connectome Project third data release.^31^ The functional images in template space were smoothed using FSL (5 mm full width half maximum gaussian kernel).^33^ This resting-state fMRI data in the common template space can be seen as forming the basis of the normative neonatal functional connectome, from which the network effect of stroke lesions can subsequently be investigated (see the following section). An independent component analysis was conducted to demonstrate the resting state networks identifiable in this cohort, which appeared as expected based on prior work already done in this cohort.^19^

#### Computation of functional connections to neonatal stroke lesions

After aligning the clinical and normative data to a common template space, it was possible to derive the functional connections to the neonatal stroke lesions. This was performed using the FSL ‘fsl_glm’ tool, where each of the stroke lesion mask images (*N*=85) were used as regions of interest in seed-based correlation analyses.^33^ Specifically, each voxel in each of the normative neonatal resting-state fMRI images (*N*=518) was correlated with the average resting-state time series signal from each stroke lesion mask region (*N*=85); i.e., each voxel in the whole brain was regressed against the lesion masks. Both positive correlations and negative correlations (anticorrelations) were possible at the individual level. The seed regions were not masked out of the analysis, meaning self-correlations (i.e., seed region correlated with seed region) were possible at the individual level (such self-correlations are typically high). This step generated a measure (*z*), which was the correlation coefficient (*r*) transformed to a z distribution using the Fisher z-transformation (equation 1):

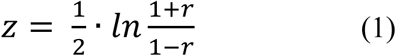

This voxel-wise measure (*z*) describes the functional connectivity between the lesions and any brain location. Thus, the output was whole-brain, voxel-wise lesion network *z* maps.^13, 21^ There were *N*=85×518 voxel-wise lesion network maps in total; one for each NAIS lesion mask region correlated with each normative resting state fMRI scan. The lesion network maps were then restricted to gray matter regions, including cortical gray matter, deep gray matter, and cerebellar regions. ^32–36^ This step was achieved through utilizing the Melbourne Children’s Regional Infant Brain atlas (M-CRIB), which provides 100 parcellated cortical gray matter, subcortical gray matter and cerebellar regions for neonatal scans.^36^ The M-CRIB structural template was registered to the developing Human Connectome Project 40-week T2-weighted template using a combination of linear warping (with FSL FLIRT^32–34^) and affine nonlinear warping (with ANTs^35^), and then the lesion network maps in template space were masked by the M-CRIB gray matter segmentation^36^ for subsequent analysis.

### Statistical analysis

Baseline participant characteristics were summarized using numbers and percentages for categorical variables or medians and interquartile ranges for continuous variables. To address the study’s aim, we tested the relationship between neonatal lesion networks and the subsequent development of cerebral palsy. Specifically, the lesion network *z* maps (*N*=85×518) were compared between participants who did and did not develop cerebral palsy after NAIS. This comparison was performed with multi-level linear regression models, to account for the interdependence of the observations (where each lesion image was correlated with the same normative images). Implementation of these models in the lme4 package^37^, R software, is shown below (equation 2):

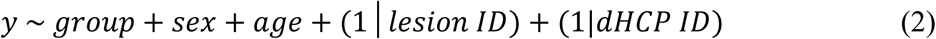

In the above, the tilde (∼) is used to separate the left-hand side (*y*) and right-hand side (group, etc.) in the model formula. The overall expression indicates that the left-hand side (the response, *y*) is modelled by a linear predictor specified on the right-hand side (group, etc.). Here, *y* represents the voxel-wise lesion network *z* map measure (as described in the section above), group represents cerebral palsy or no cerebral palsy, sex represents participants’ sex, age represents participants’ age at MRI, and lesion ID and developing Human Connectome Project (dHCP) ID represent random effects to account for repeated observations within participants.

The output was a voxel-wise lesion network *contrast* map (i.e., from contrasting the lesion network maps between those with and without cerebral palsy, adjusted for age and sex). These voxel-wise lesion network contrast maps were *t*-statistic maps, which were corrected for voxel-wise multiple comparisons using the Statistical Parametric Mapping software package (SPM12).^38^

Significant gray matter regions detected by this analysis were more highly functionally correlated with lesions in participants who developed cerebral palsy compared with participants who did not develop cerebral palsy; i.e., this identified gray matter regions part of functional networks significantly related to the development of cerebral palsy following NAIS. Statistical significance was defined as *t*>5.4 and *p*<0.05, family-wise error rate (FWE)-corrected. The subsequent results section focusses on reporting the range of *t*-values, noting all *p*-values were less than 0.05, FWE-corrected.

## Results

### Participant characteristics

From the total 199 participants recruited into the Australian and Swiss pediatric stroke registries, a subset of 85 newborns with NAIS were included in the current study. Exclusions (*N*=114) were due to loss to follow-up, unavailable or poor-quality imaging, subacute infarction, dual pathology, and lesion segmentation failure, as determined by experienced pediatric neurologists. Key characteristics that are relevant to the current analysis of the 85 newborns with NAIS are presented in Table 1. As shown, the median age at stroke symptom onset for the included NAIS participants was 2 days (interquartile range 1–2 days), and the median age at MRI scanning for the included NAIS participants was 4 days (interquartile range 3–5 days). All NAIS participants underwent MRI within 10 days of stroke symptom onset. All characteristics shown did not differ between participants with and without cerebral palsy (all *p*>0.05), except lesion volume was higher in those with cerebral palsy (median: 52, interquartile range: 23-113 cm^3^) compared to those without cerebral palsy (median: 13, interquartile range: 7-29 cm^3^; *p*<0.001). From examining the distribution of lesion volume data, approximately one third of participants in the cerebral palsy group had lesion volumes that fell below or within the interquartile range of lesion volumes in the group without cerebral palsy. This highlights that in some participants with cerebral palsy, lesion volume is not high, and thus lesion volume is not a strong predictor of cerebral palsy for all cases (as noted in prior work).^5^ Additionally, all characteristics shown did not differ between participants from Australia and Switzerland (all *p*>0.05). From the total 783 participants provided by the developing Human Connectome Project’s third open access data release, we used data for 518 term-born newborns. Exclusions (*N*=265) were due to preterm birth or a lack high-quality structural and resting state fMRI data. Key characteristics of the 518 normative participants are presented in Table 2.

**Table 1.** Neonatal stroke patient characteristics.

| Characteristic | Neonatal stroke patients (N=85) |
| --- | --- |
| Australian, N (%) | 51 (60) |
| Male, N (%) | 55 (64) |
| Gestational age at birth (weeks), median (IQR)<sup>a</sup> | 40 (39-41) |
| Birth weight (kg), median (IQR)<sup>b</sup> | 3.4 (3.1-3.7) |
| Age at stroke (days), median (IQR) | 2 (1-2) |
| Age at MRI (days), median (IQR) | 4 (3-5) |
| Lesion volume (cm<sup>3</sup>), median (IQR) | 22.3 (10.5-49.2) |
| Supratentorial brain volume (cm<sup>3</sup>), median (IQR) | 398 (382-424) |
| Age at follow-up (years), median (IQR) | 2.1 (1.9-2.6) |
| Cerebral palsy, N (%) | 28 (33) |

IQR = interquartile range; N = number. <sup>a</sup>N=81; <sup>b</sup>N=82.

**Table 2.** Developing Human Connectome Project participant characteristics. Characteristic Normative participants (*N* = 518)

| Characteristic | Normative participants (N = 518) |
| --- | --- |
| Age at birth (weeks), median (IQR) | 40 (39-41) |
| Birth weight (kg), median (IQR) | 3 (3-4) |
| Male, *N* (%) | 280 (54) |
| Singleton, *N* (%) | 498 (96) |

IQR = interquartile range; N = number.

### Neonatal lesion networks and cerebral palsy

Results of the neonatal lesion network mapping analysis are shown in Fig 2 and Table 3. Fig 2 visually displays the significant regions in the neonatal lesion network contrast maps, and Table 3 lists all significant regions by brain hemisphere (left or right). As shown in Fig 2 and Table 3, in participants who developed cerebral palsy compared with participants who did not develop cerebral palsy after NAIS, lesions were more highly functionally connected to several cortical, cerebellar and deep gray matter regions. This included the following: basal ganglia (caudate, putamen, pallidum, accumbens), thalamus, cerebellum, frontal (inferior and orbital frontal, superior frontal), temporal (pole, superior, and mesial temporal including hippocampus and amygdala) and insula (Fig 2, Table 3).

**Fig 2.**
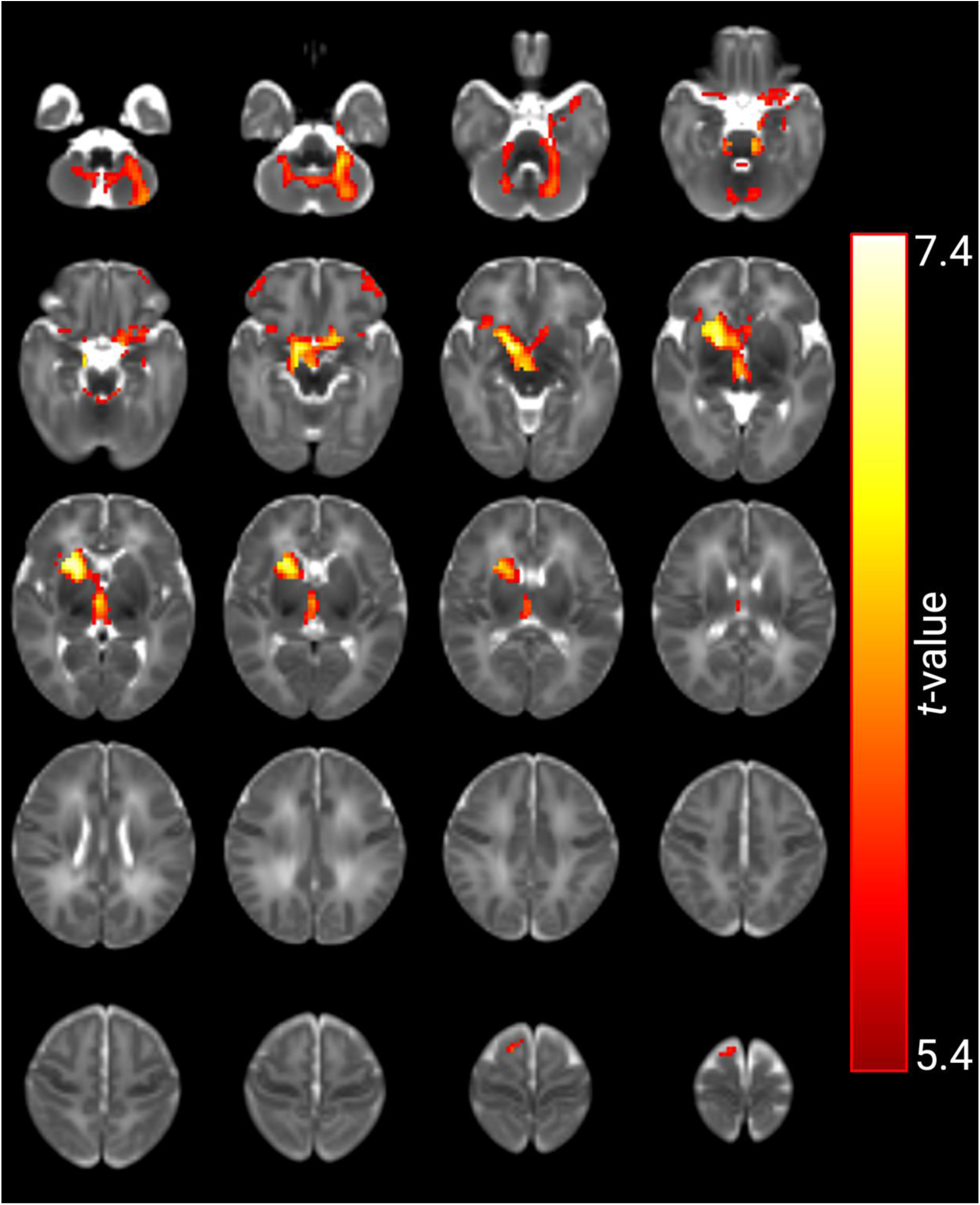
Results of the neonatal lesion network mapping approach: the neonatal lesion network contrast maps. The brain regions highlighted in the red-yellow color scale are all statistically significant (*t* > 5.4; *p* < 0.05, family wise error rate-corrected), where increasingly yellow colors indicate the regions were more highly functionally connected to lesions in those who developed cerebral palsy compared with those who did not develop cerebral palsy. Results are overlaid on the developing Human Connectome Project 40-week T2-weighted template image. Images are presented in radiological convention, where the left hemisphere of the brain is shown on the right side of the image. A full list of regions is provided in Table 3.

**Table 3.** Complete list of significant brain gray matter regions identified in the neonatal lesion network mapping analysis (i.e., in the lesion network contrast maps).

| **Brain region** | **Maximum *t*-value** |
| --- | --- |
| Right pallidum | 7.35 |
| Right putamen | 7.20 |
| Right amygdala | 7.06 |
| Right caudate | 7.00 |
| Brainstem | 6.61 |
| Right thalamus | 6.55 |
| Right hippocampus | 6.54 |
| Left cerebellar hemisphere | 6.51 |
| Left lateral orbitofrontal | 6.44 |
| Left accumbens | 6.35 |
| Right cerebellar hemisphere | 6.31 |
| Right lateral orbitofrontal | 6.06 |
| Left thalamus | 5.92 |
| Left putamen | 5.89 |
| Left entorhinal | 5.88 |
| Left insula | 5.86 |
| Cerebellar vermis inferior posterior | 5.86 |
| Right insula | 5.84 |
| Right superior frontal | 5.79 |
| Left medial orbitofrontal | 5.75 |
| Left temporal pole | 5.72 |
| Left amygdala | 5.66 |
| Right medial orbitofrontal | 5.65 |
| Right accumbens | 5.63 |
| Left caudate | 5.63 |
| Cerebellar vermis anterior | 5.58 |
| Left superior temporal | 5.54 |
| Right pars orbitalis | 5.52 |
| Left hippocampus | 5.46 |
| Left rostral anterior cingulate | 5.44 |
| Right rostral anterior cingulate | 5.38 |
| Left parahippocampal | 5.37 |

### Further analysis of neonatal lesion networks

To further understand the results in the prior section, we extracted average lesion network *z* values from the significant brain regions in the contrast maps. To do this, we started from the *N*=85×518 voxel-wise lesion network *z* maps. For each of the 85 newborn stroke participants, we averaged the respective 518 voxel-wise lesion network *z* maps, resulting in 85 voxel-wise lesion network *z* maps. For each of these 85 maps, we averaged the *z* values within each significant region (i.e., each region identified in the prior section). The average lesion network *z* values for example significant cerebellar, cortical and deep gray matter regions are shown in Fig 3.

**Fig 3.**
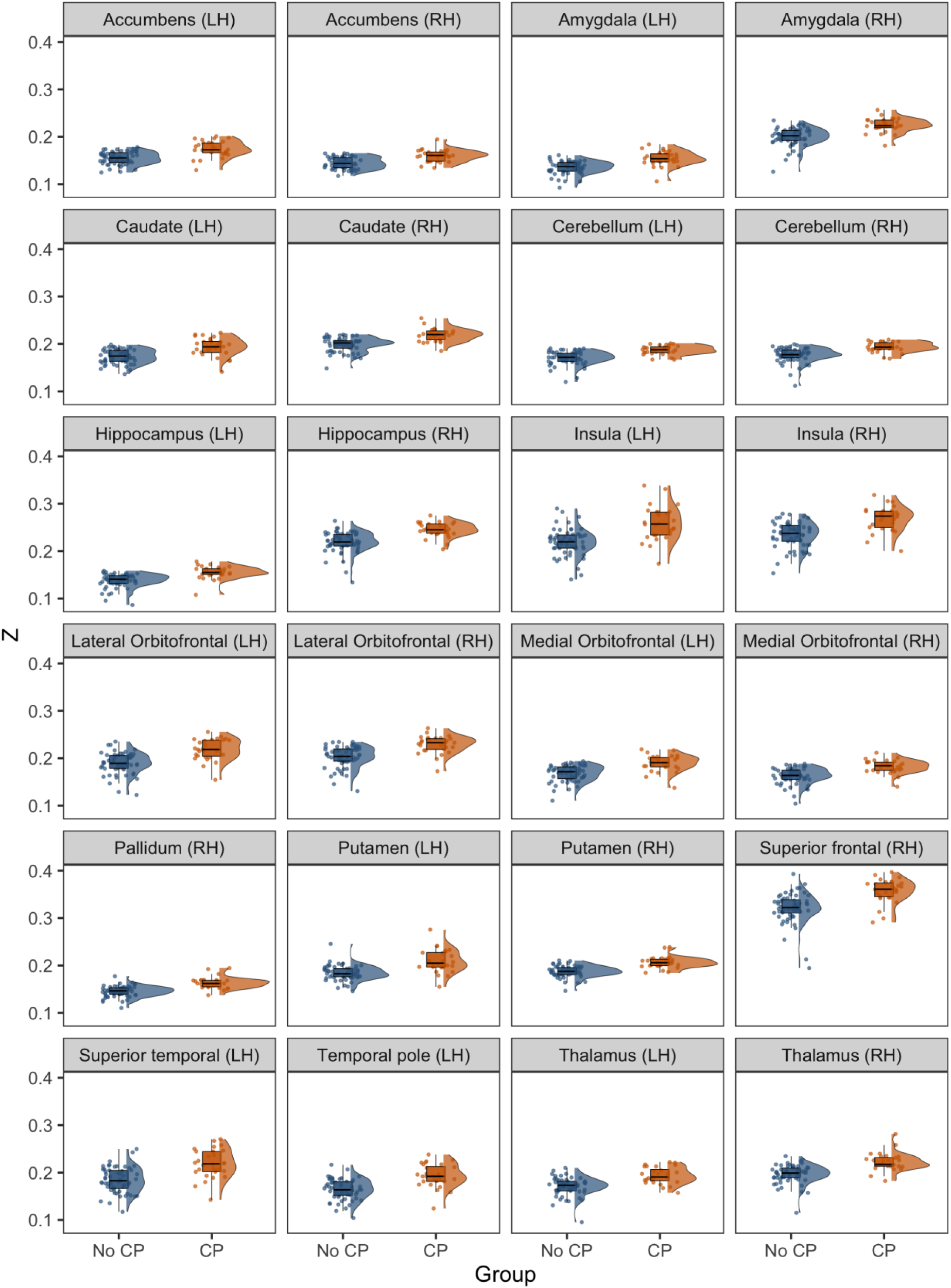
Neonatal lesion network *z* maps. In this figure, the *y* axis is the average neonatal lesion network *z* measure for each region shown [where *Z* = correlation coefficient (for the functional correlation between lesions and other brain regions) transformed to a z distribution, as described more in the methods section of the current manuscript]. This average lesion network *z* measure has been presented in the form of raincloud plots (jittered raw data, box plots, and split violin plots), separately for the participants with and without cerebral palsy (CP). This figure demonstrates that functional correlations between lesions and other brain regions were positive correlations, with no negative correlations observed. Furthermore, the positive correlations were greater in magnitude in the participants who developed CP compared with the participants who did not develop CP.

Average lesion network *z* values in significant brain regions were both positive, and greater in magnitude in the participants who developed cerebral palsy compared with the participants who did not develop cerebral palsy (Fig 3). This indicates that, while both positive correlations and negative correlations (between the neonatal stroke lesions and the rest of the brain gray matter) were possible on an individual level, only positive correlations were identified as significant at a group level in the current study.

## Discussion

This study developed a neonatal stroke connectome, through leveraging clinically acquired acute diffusion imaging in combination with a large-scale resting state fMRI dataset. This led to the identification of multiple deep gray matter, cortical, and cerebellar regions associated with the development of cerebral palsy. These regions included some that are not typically implicated in adverse motor outcomes.

Within the functional connectome, the gray matter regions most related to the development of cerebral palsy after neonatal stroke were the deep gray matter regions, including the caudate, putamen, pallidum, accumbens, and thalamus, and the cerebellum. The regions next most implicated were the frontal regions (inferior, orbital frontal and superior frontal), temporal regions (pole, superior, and mesial temporal including hippocampus and amygdala) and insula. When comparing our results with prior imaging studies of NAIS cohorts, we believe our findings are both complimentary and unique, in that there is overlap and differences in the regions found in the current study and regions found in prior studies.^5–8^ The most relevant prior studies are those that utilized the VLSM approach.^5–7^ VLSM and lesion network mapping are similar in that they survey much of the brain on a voxel-wise basis. However, prior VLSM approaches focused on surveying lesion regions (encompassing lesions in both gray and white matter), whereas the current lesion network mapping approach is capable of surveying gray matter regions remote from lesions via their functional connections. These methodological similarities and differences could account for some similarities and differences in results observed in the current study and prior VLSM studies. Summarizing prior studies, Nunez *et al.*^6^ found that NAIS lesions in white matter regions (the external capsule and corona radiata) correlated with motor function impairment at age 2 years. Dinomais *et al.*^7^ found that NAIS lesions in white matter regions (the junction of the superior longitudinal fasciculus and corona radiata) and gray matter regions (insula, precentral gyrus and postcentral gyrus) correlated with unilateral cerebral palsy. Both the current study and our prior VLSM study of the same NAIS cohort by Mackay *et al.*^5^ found that deep gray matter regions (putamen, pallidum, caudate, accumbens, thalamus), insula, frontal regions (superior frontal and orbital frontal), and temporal regions (superior temporal and mesial temporal including hippocampus and amygdala) were related to cerebral palsy. In this prior VLSM study^5^ but not in our current study, the precentral and postcentral cortical regions were related to cerebral palsy. Conversely, in our current study but not in our prior study by Mackay *et al.*,^5^ the cerebellar regions were related to cerebral palsy.

To summarize, the deep gray matter regions have been commonly identified as related to motor outcomes in both the current study and prior studies. However, the current study found that the extramotor frontal and temporal cortical regions were related to the development of cerebral palsy, while prior studies found that primary motor cortical regions were implicated in the development of cerebral palsy (including the precentral and postcentral cortical regions, which were not significantly associated with cerebral palsy in the current study).^5–8^ Some of the gray matter regions found in the current study have previously been implicated in motor function, including the basal ganglia as part of the extra-pyramidal cortical-striatal motor pathway, the thalamus as part of the somatosensory pathway, the cerebellum, the insula as part of the primary motor pathway involved in motor control, and the superior frontal regions as part of the association motor cortices.^5^ However, some of the gray matter regions found in the current study have not previously typically been related to motor outcomes, such as the temporal regions (pole, superior, and mesial temporal including hippocampus and amygdala) and inferior, orbital frontal cortical regions.^5^ Possible explanations for these findings are discussed in the following paragraphs.

This study using a novel connectome-based approach has identified extramotor regions that are functionally connected to NAIS lesions, and where this functional connection relates to the development of cerebral palsy. This finding could represent broader network disruptions following NAIS. Network disruption is supported by the identification in our results of centrally located, highly connected structures like the thalamus,^14^ and by the identification in our results of the cerebellum, where we know that stroke does not occur within the cerebellum in NAIS.^5^ We found regions that overlap with prior lesion maps^5^, along with regions that are outside known NAIS lesions (e.g., the cerebellum), meaning our results are unlikely to reflect lesion extension or new areas of infarction within extramotor regions following NAIS. We believe our findings support the concept of functional disconnection or diaschisis and pre-Wallerian degeneration following NAIS, which have been suggested to occur in prior studies.^13, 14, 39^ Generally, these phenomena have been seen as established in NAIS cohorts for primary motor regions and association motor regions, but not necessarily for some of the temporal and inferior, orbital frontal cortical regions found in the current study; however, our study suggests these additional regions could be involved.^13, 14, 39^

Prior studies have established that resting-state networks are already present in newborns as early as term-equivalent age,^19^ and that motor outcomes in healthy populations and other clinical populations are related to both primary motor networks and other broader motor networks.^40, 41^ Our observation that the development of cerebral palsy correlated with both motor and extramotor regions lines up with these findings in other populations, and may indicate more widespread network vulnerabilities after stroke in neonates compared with adults.^21^ The extramotor regions (the frontal and temporal regions identified in the current study) may be more vulnerable to network disruptions in neonatal stroke due to the known relative immaturity of these regions during the neonatal period and their prolonged maturation to support higher-order functions.^42, 43^ Particular vulnerability of these temporal and frontal regions has been demonstrated in large-scale, longitudinal studies of other newborn, very preterm populations at risk of brain insult and injury.^43^

There are limitations to this study. Lesion network mapping has frequently been performed in adults, and is more clinically feasible than directly acquiring resting-state fMRI scans in NAIS patients, however lesion network mapping may be seen as a less direct method for assessing brain connectivity.^13, 21, 22^ Functional connectivity has been well characterized in adults and neonates,^1, 16–19^ but future studies will need to examine structural connectivity between regions via white matter fiber pathways using diffusion MRI scans to determine whether the functional changes identified correspond to structural changes.^44^ Given the paucity of prior research in this area, we aimed to establish that there is a relationship between the lesion functional connectome and cerebral palsy, but future work will also need to investigate the relative contributions of different factors (such as lesion volume, lesion location, and lesion functional and structural connectivity) to the development of cerebral palsy. Given this study utilized clinically indicated MRIs from multiple sites for stroke participants, uniform scanner (e.g., field strength) and sequence (e.g., resolution) settings could not be used. The lesion network mapping approach does not necessarily require uniform scanning, making this approach more accessible. However, we acknowledge that variations in scanning parameters could influence some analysis steps, such as lesion segmentation and mapping to the template. To account for this, all analysis steps including lesion segmentation and registration were performed by experienced pediatric neurologists and image scientists and outputs were extensively visually checked to ensure any poor-quality data were excluded.

Lesion network mapping identifies correlations between lesions and the rest of the gray matter, which could include the lesion regions themselves (self-correlations, which are typically likely to be high). If self-correlations were high in both groups (the cerebral palsy and non-cerebral palsy groups), then no significant group differences would be identified, which could explain why the primary motor regions did not appear in the current findings (as the primary motor regions are key lesion regions affected by NAIS). The correlations identified between lesions and gray matter regions were positive correlations (rather than anti-correlations), which could reflect increased activity to support motor function (as opposed to decreased activity of irrelevant processes), as seen during task performance but even intrinsically occurring at rest.^45^ We also note that positive correlations are in general stronger and less variable than anti-correlations.^46^ We also identified significant regions in both the left and right hemispheres of the brain; however, future work to specifically test whether there was a difference in the findings between hemispheres would be worthwhile (i.e., to test whether results were greater in magnitude or spatial extent in the left or right hemisphere, which could be related to asymmetry in the lesions or connectome or both). With lesion network mapping, it was possible for us to infer that the regional networks identified in our study correlated with cerebral palsy, but we cannot necessarily extrapolate on the relative importance of the individual regions of the network in cerebral palsy based on this method alone.^13^

Other limitations include challenges with case ascertainment, with data being available for less than half of eligible children recruited to the pediatric stroke registries.^5^ Clinical diagnosis of cerebral palsy was determined at age 2 years, while follow up at age 4 years may be preferred to avoid misdiagnosis or missing milder cases of cerebral palsy.^5^ Acknowledging that many newborns with NAIS exhibit both motor impairments and impairments in a range of other domains including cognition, language and behaviour,^47, 48^ further work to predict these additional outcomes based on MRI data will be important.

In conclusion, this study identified novel acute connectome-based correlates of cerebral palsy following neonatal stroke. The findings could suggest that development of cerebral palsy after neonatal stroke is not solely based on damage to primary motor systems but is related to the involvement of broader networks. This has important implications for researchers, clinicians and families by highlighting that when predicting the development of cerebral palsy following neonatal stroke, it is important to consider regions that are supportive of, but not directly included in, the primary motor system. In future, the ultimate goal could be to rapidly produce multi-modal connectomes (utilizing ongoing technological developments such as automated lesion segmentation and deep learning techniques for connectome production^49^) and predict long-term motor outcomes for each individual neonate with stroke, to guide personalized neonatal care by impacting the way each baby is monitored and referred to intervention services.

## Non-standard Abbreviations and Acronyms

ANTs: Advanced Normalization Tools
AP: anterior-posterior
CP: cerebral palsy
dHCP: developing Human Connectome Project
FLIRT: The FSL Linear Image Registration Tool
fMRI: functional MRI
FSL: The Functional MRI of the Brain Software Library
FWE: family-wise error rate-corrected
ICA: independent component analysis
IQR: interquartile range
M-CRIB: Melbourne Children’s Regional Infant Brain atlas
NAIS: neonatal arterial ischemic stroke
PA: posterior-anterior
SENSE: sensitivity encoding
SPM12: Statistical Parametric Mapping
TE: echo time
TR: repetition time
VLSM: voxel-based lesion-symptom mapping.

## Acknowledgements

This research was supported by The Royal Children’s Hospital Foundation, Murdoch Children’s Research Institute, The University of Melbourne, Department of Paediatrics, and the Victorian Government’s Operational Infrastructure Support Program. We would like to thank the staff of the individual centers of the Swiss Neuropediatric Stroke Registry and Dr Maria Regenyi from Bern, who helped with data collection. We are grateful to the families who generously supported the developing Human Connectome Project.

